# Prevalence of Body-focused repetitive behaviours among undergraduate medical students at public sector university karachi

**DOI:** 10.1101/2024.08.20.24312259

**Authors:** Marium Munir Tunio, Eshika, Vandna Devi, JaiRam, Kirshna, Sandesh, Vandna Kumari, Jamshaid

## Abstract

**Background:** Body-focused repetitive behaviors (BFRBs) is a catch-all term for a group of obsessive behaviors, including skin picking (dermatillomania), hair pulling (trichotillomania), nail-biting (onychophagia), and nose picking (rhinotillexomania), which have a negative impact on both physical and mental health. The aim of this study was to assess the prevalence of BFRBs among medical students aged 18 to 25 attending Jinnah Sindh Medical University Karachi.

**Methodology:** A cross-sectional study was conducted on 295 medical students of Jinnah Sindh Medical University Karachi. Data was collected through an online questionnaire which consisted of socio-demographic characteristics, engagement in BFRBs and distress due to it.

**Results:** Majority of participants (61.2%) were in their clinical years (3rd, 4th and 5th). Among those exhibiting BFRBs, 69.9% were female. Through this study we found that the most common BFRB among the medical students of Jinnah Sindh Medical University is Nail biting which is 22.8% followed by Skin picking disorder which is 19.1%,Hair pulling 17.1%,Cheek biting 13.8% followed by Nose picking 7.7%

**Conclusion:** In our research, it has been noticed that participants were engaged in BFRBs. But it was not significantly common in adults. Majority of respondents reported never engaging in these behaviors (skin picking, hair pulling, nail-biting, nose picking and cheek biting). While a significant percentage of respondents engaged occasionally or frequently in these behaviors.

## INTRODUCTION

Body-focused repetitive behaviors (BFRBs) is a catch-all term for a group of obsessive behaviors, including skin picking (dermatillomania), hair pulling (trichotillomania), nail-biting (onychophagia), and nose picking (rhinotillexomania), which have a negative impact on both physical and mental health.[1]

The application of BFRBs is to ease or alleviate stress-related negative emotional states, such as a link with states and trait anxiety. When they occur less frequently, these illnesses are generally benign (subclinical), Yet, when these BFRBs reach this threshold, they are categorized as impulse-controlled diseases (pathological) [2]. Treatment may be necessary if a person’s actions seriously impair their quality of life or show signs of extreme anxiety.

This pathogenic threshold is not usually met in many cases[3]. Yet when it develops, therapies such as habit reversal training and decoupling, which are used to treat other BFRBs, can be used.[4]

One such condition is nail biting (NB) in which a patient shows self-harming potential, due to surrounding stressful and social conditions [5]The term “nail-biting” (NB) describes the insertion of the fingers into the mouth and the subsequent contact of the nails with the teeth. NB as a BFRB entails biting past the nail bed and cuticles, drawing blood, and resulting in persistent scarring, or in red, painful, and infected fingers. It is common for people to use their teeth as an occasional replacement for nail clippers when grooming themselves.[6]

Hair pulling also known as (trichotillomania) characterized by the chronic and repetitive pulling of hair, which can lead to visible hair loss. People with this disorder often make repeated attempts to stop or reduce hair-pulling but are unable to do so, causing significant distress and impairment. Hair pulling can affect any part of the body where hair grows, including the scalp, eyebrows, arms, legs, and pubic area, as stated by the American psychiatrist Association [7]. Triggers to pull include sensory (i.e., hair thickness and length), emotional (i.e., feeling anxious), and cognitive (i.e., thoughts about hair and appearance)cues [8] Psychosocial dysfunction, low self-esteem, and social anxiety are all associated with hair pulling, largely due to an inability to stop pulling and the resulting alopecia[9][10].

Skin excoriation is an other term for (dermatillomania), commonly known as skin-picking disorder (SPD). It alludes to repeated attempts to lessen or halt Skin-Picking (SP), which causes sores. The symptoms are not better described by another mental condition and produce clinically considerable distress, and impairment in social, occupational, and other key areas of functioning [11]. For most individuals, picking, squeezing, or scratching to get rid of pimples, crusts, or other skin blemishes is a regular cosmetic regimen. A significant segment of the populace often admits to occasionally or frequently participating in this skin-picking behaviour.[12].

The practice of picking one’s nose involves removing nasal mucus with one’s finger (rhinotillexis), which may also involve swallowing the removed mucus (mucophagy)[13].Rhinotillexomania is the term for nose picking that develops into an obsessive-compulsive disorder or repetitive body-focused behaviour. Themajority of instance fall short of this pathological standard [14[15] [16][17]. Yet when it develops, therapies such as habit reversal training and decoupling, which are used for other BFRBs, can be used.

Cheek biting (morsciato bacurum) is a disease marked by chronic buccal mucosa irritation or damage brought on by frequent chewing, biting, or nibbling [18]. The majority of people who accidentally bite their cheeks don’t experience any harm, but those who have made it a practice of doing so may experience harm.

The prevalent causes of morsicatio buccarum include ill-fitting dentures, jaw closure issues, temporomandibular joint disorders, and misaligned wisdom teeth. However, morsicatio buccarum is frequently seen in individuals who are anxious, stressed, or have a psychogenic background [19]

### RATIONALE

For a number of reasons, it is crucial to do research on the prevalence of BFRBs among medical students. First off, because of the high amounts of stress and pressure connected with a medical school, medical students may be more likely to acquire BFRBs, which can have a severe effect on their job prospects and psychological well-being. Finally, studies on the prevalence of BFRBs among medical students can help with the creation of focused therapies and support systems. To minimize BFRBs and lessen their influence on everyday functioning, medical students may benefit from stress management programmers or cognitive-behavioral therapy.

### METHODOLOGY

Study setting and Participants study will be conducted in Sindh medical college Karachi. The medical school has a diverse student body, with students from various ethnic, cultural, and socioeconomic backgrounds. The participants in this study will be medical students who are currently enrolled in MBBS program of SMC.

#### Research Design

Study design for this study is a cross-sectional survey.In this case, the population of interest is medical students of SMC and the survey will be used to assess the prevalence of bodyfocused repetitive behaviours (BFRBs) in this population.

#### Strategies to Minimize Bias

In this study, various measures were implemented to minimize potential biases. The sampling was conducted using randomized selection to reduce selection bias and ensure a representative sample of the target population. Data collection was standardized through the use of validated self-report questionnaires. To mitigate recall and reporting bias, participants were provided with clear instructions and reassured of confidentiality to encourage honest and accurate responses. Additionally, statistical methods were applied to adjust for potential confounders and reduce their influence on the study outcomes

#### Sample Size

The study sample size was calculated through online available OpenEpi calculator. The following assumptions were made to calculate the sample size:

n= (Zα/2)2 (pq) /(B)2 n= (1.96)2 (0.22*0.78)/ (0.05)2 n=263.66

Precision = 5%

Prevalence= 22% [20]

95% Confidence Interval

The sample size came out to be 263.66 which is rounded off to 264 this means that a minimum of 264 participants would be required to estimate prevalence of BFRBs with a margin of error of +/-5% and confidence interval of 95% after adjustment for 10% non response rate sample size is inflated to 293

#### Sampling Technique

Non-probability convenience sampling technique will be used to recruit students enrolled in MBBS program at Sindh medical college.

#### Inclusion Criteria

- Currently enrolled in MBBS program, Male and female medical university students, willing to give informed consent.
- Age between 18 years to 25 years.
- Those who consent to be a part of the study

#### Exclusion Criteria

- Not currently enrolled in the MBBS program of Sindh medical college.
- Unable or unwilling to provide informed consent to participate in the study.
- Students aged below 18 and above 25.

#### Data Collection Tools

- Self-administered structured questionnaire (consisting of 26 questions) which has been constructed after a thorough review of literature will be used to obtain information regarding socio-demographic characteristics and personal

characteristics of the study participants and to figure out whether they experience BFRBs and calculate its prevalence among our study participants.

#### Data collection

The questionnaire will be tested through a pilot study prior to data collection.

#### Study Analysis

The descriptive parameters of the student’s data were determined in terms of frequencies, along with the types of Body Focused Repetitive habits (Bfrbs), their effects, prevalence, and techniques for quitting these habits. Values for the means, medians, and modes were also computed. To provide a clear and comparative picture, some data is presented as “pie charts”. Odds ratios with a 95% confidence interval, a 5% margin of error, and a 0.05 p-value were considered statistically significant.

“SPSS Version 20” was used to analyse the data.

## Results

All 299 students invited to participate in the study completed the survey. There were no instances of non-participation.

Out of total 299 students 45.59%(n=136) were 21-22 years old with females 69.9%(n=209) and 61.2%(n=183) in clinical years of study (Table 1)

**Table 1:** Socio -demographic characteristics of study participants (n=299)

|  | % of students | No of students(n) |
| --- | --- | --- |
| Age groups |  |  |
| 18-20 | 31.7% | 111 |
| 21-22 | 45.5% | 136 |
| 23-25 | 16.1 % | 48 |
| Gender |  |  |
| Male | 28.8% | 86 |
| Female | 69.9 % | 209 |
| University |  |  |
| JSMU | 100 % | 299 |
| Study years |  |  |
| <u>Non- clinical(1st and 2nd year)</u> | 37.5% | <u>112</u> |
| Clinical(3rd,4th and5th year) | 61.2% | 183 |

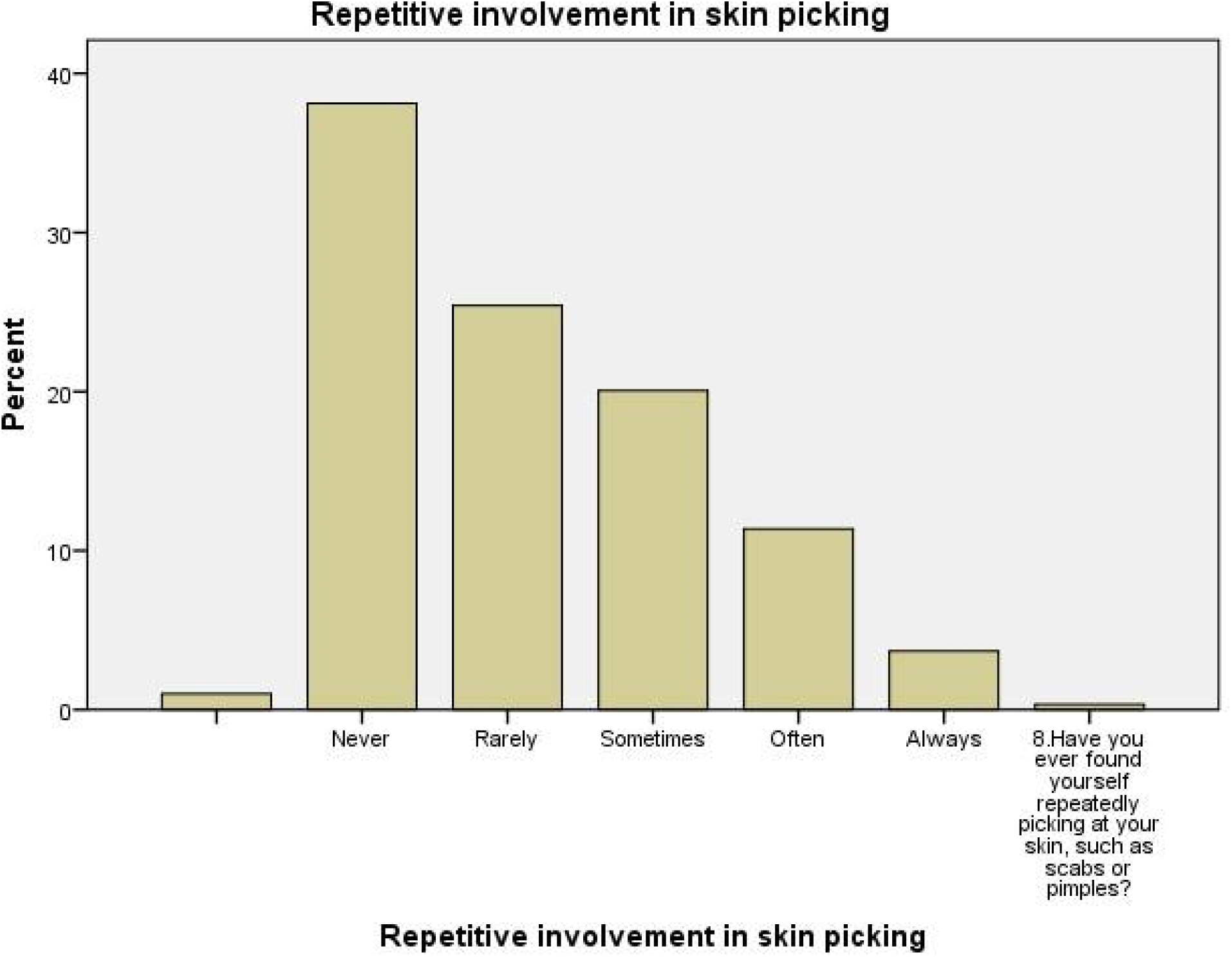

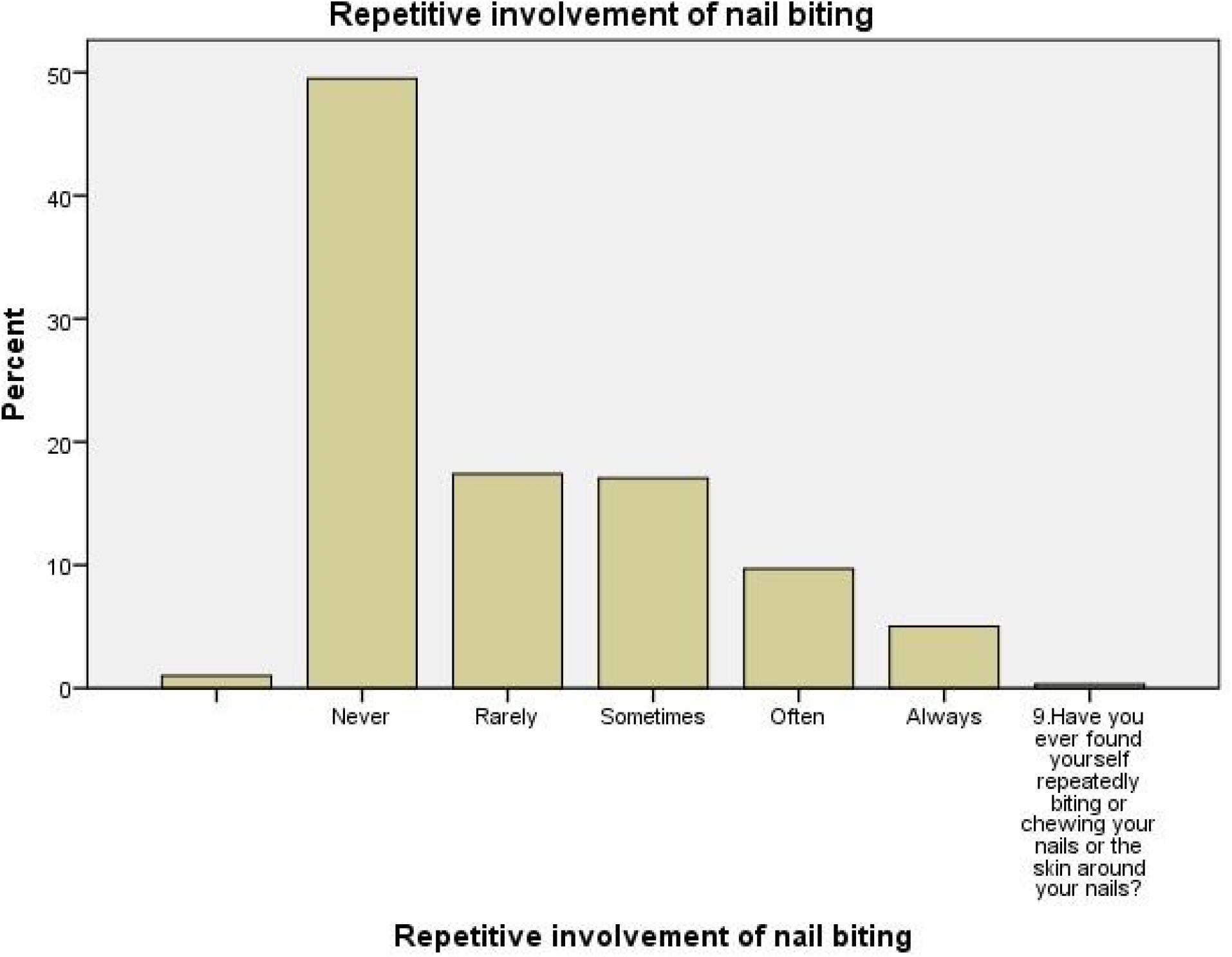

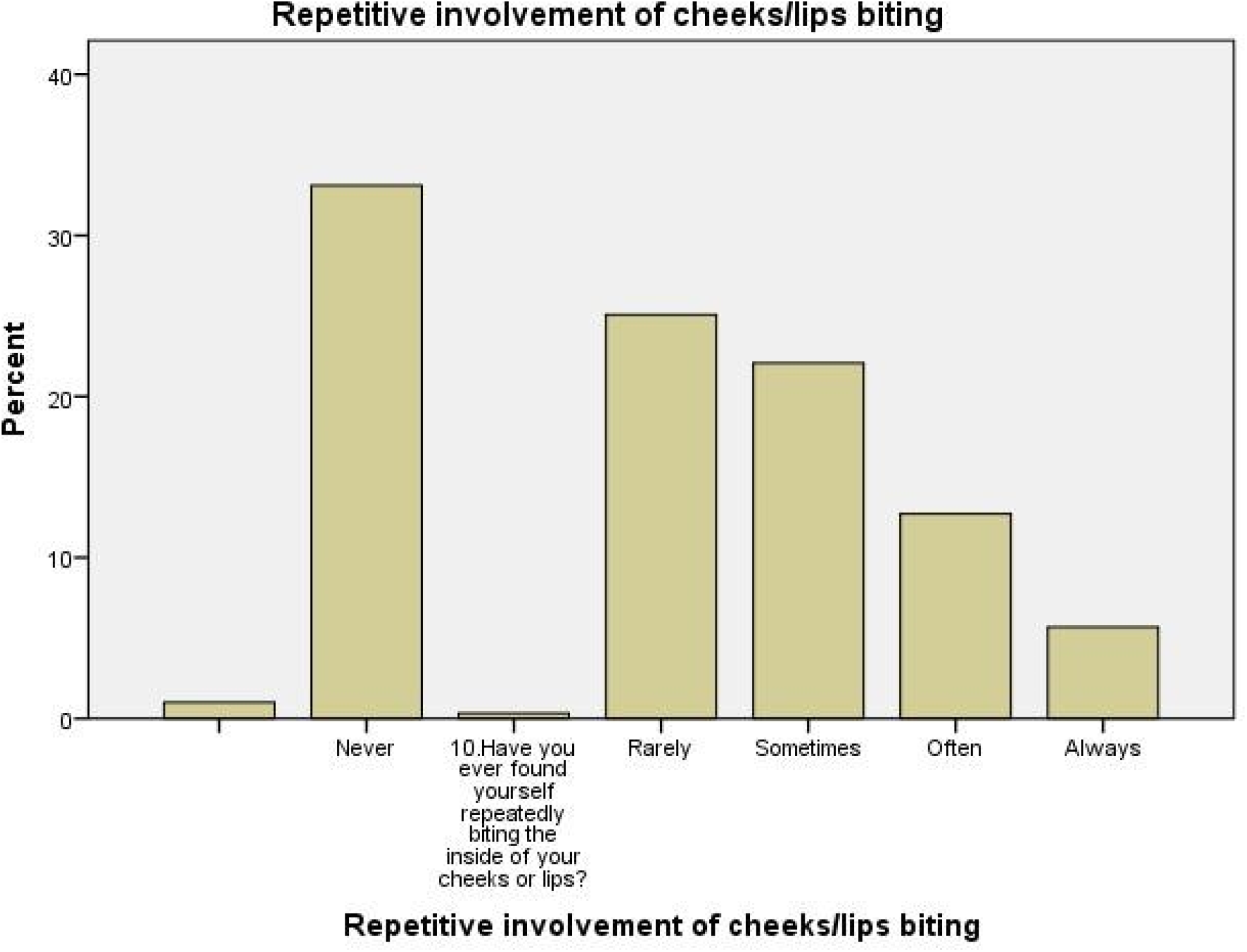

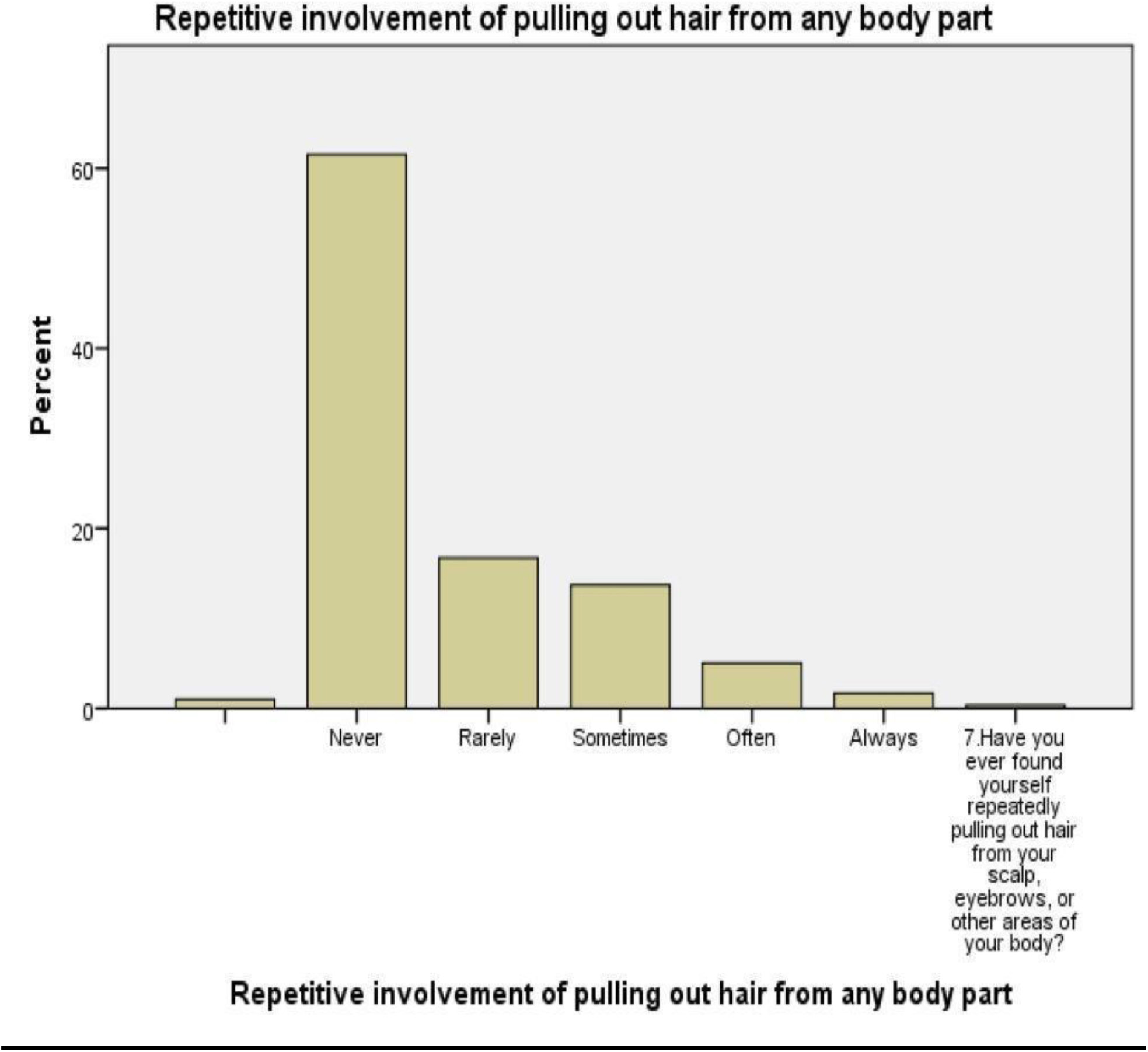

A significant 61.5% of 299 respondents never engage in repetitive hair removal, while 16.7% rarely, 13.7% sometimes, 5.0% often, and 1.7% always.Significant p value <0.001.

38.1% of respondents never engage in repetitive skin picking, while 25.4% rarely, 20.1% sometimes, 11.4% often, and 3.7% always.Significant p value <0.001.

49.5% of 299 participants never engage in nail biting, while 17.4% rarely, 17.1% sometimes, 9.7% often, and 5% always engage in this behavior.Significant p value <0.001.

33.1% of 299 respondents never experienced repetitive involvement of cheeks/lips biting, while

0.3% rarely, 25.1%sometimes,12.7%often,and 5.7% always.Significant p value <0.001. 55.5% of respondents never extract nasal mucus, with 0.3% rarely, 24.1% sometimes, 4.3% often, and 0.7% always.Significant p value <0.001.

## Conclusion

In our research, it has been noticed that participants were engaged in BFRBs. But it was not significantly common in adults. Majority of respondents reported never engaging in these behaviors (skin picking, hair pulling, nail-biting, nose picking and cheek biting). While a small percentage of respondents engaged occasionally or frequently in these behaviors. The research also found that these behaviors more common in 21-22 age group, clinical years (3rd,4th and 5th) and females.

## Discussion

BFRBs are common among medical students. Consequentially, it is crucial to assess the prevalence of BFRBs in order to develop interventions that can alleviate the impact.

Through this study we found that the most common BFRB among the medical students of Jinnah Sindh Medical University is Nail biting which is 22.8% followed by Skin picking disorder which is 19.1%,Hair pulling 17.1%,Cheek biting 1.8% followed by Nose picking 7.7%. Another study also proved Nail biting as the most common repetitive habit (Body focused repetitive behavior problems_Teng EJ, Woods DW, Twohig MP, Marcks BA)

Some studies also suggest chewing on lips as the most common repetitive habit (Research by medical students of Peshawar Medical College Disorder specific rates in our study ranged from 19.1%-22.8% for skin picking and nail biting which is higher than a previous study on medical students of Karachi which shows the rates of 9.0% and 6.2% for skin picking and nail biting.This study also suggest Hair pulling as the most common repetitive behavior 13.3%.

Research suggests that SKIN PICKING occurs in people with age 13-15 but in our study the medical students are mostly above 18 to 25 with the mean age of 21.5 years and because of our cross sectional study with limited sample it is not possible to reach valid conclusion. Among our study participants. We observed a higher prevalence among females 69.9% and 30.1%among males coherent with previous studies in which female rate is 77.5 %.

The literature indicates that stressful conditions can lead to BFRBs,but there may not be a direct causal relationship between the two. The elevated occurrence of BFRBs among females may be linked to their heightened vulnerability to stressful condition, potentially leading to more frequent engagement in these behaviors compared to males.

There are various triggers and causes of BFRBs but mostly common in all of them are stress anxiety and psychogenic background besides other reasons such as body dysmorphic disorder and, low selfesteem.Thus our study suggest that body repetitive behaviors are frequently displayed by medical students. Nail biting is the the most common repetitive habit in both males and females but women are far more likely to engage in it.Given these results additional research in the fields of neurobiology, psychiatry and epidemiology is required to understand the causes and mechanisms of BFRB and its relation with other disorders.

## Supporting information

Consent form

abstract

IRB approval

## Data Availability

All data produced in the present work are contained in the manuscript

## Limitations

The study has several limitations. First, the cross-sectional design does not allow for causality to be established. Second, the reliance on self-reported data may introduce response bias, as students may underreport or overreport their engagement in BFRBs. Finally, the study was conducted at a single institution, which may limit the generalizability of the findings to other populations. The findings of this study may be generalized to populations similar in demographic and cultural characteristics to the study participants. However, caution should be exercised when extrapolating the results to broader populations, as the sample was drawn from a specific geographic area and primarily consisted of individuals who volunteered to participate. Future studies involving more diverse populations across different regions and age groups are recommended to further validate the findings and enhance their generalizability.”

## Funding

This study did not receive any specific grant from funding agencies in the public, commercial, or not-for-profit sectors.

## Conflict of Interest

The authors declare that they have no conflicts of interest.

## Notes

### Competing Interest Statement

The authors have declared no competing interest.

### Funding Statement

This study did not receive any funding

### Author Declarations

IRB of Jinnah Sindh Medical university Karachi gave ethical approval for this work

