## Supplementary material for "Prevalence of Body-focused repetitive behaviours among undergraduate medical students at public sector university karachi": Consent form

**Project Information**

| Project: Prevalence of Body-focused repetitive behaviors among undergraduate medical students at a public sector university karachi |  |
| --- | --- |
| IRB Ref No: | JSMU/IRB/2023/749 |
| Principal Investigator: Dr. Marium Munir Tunio | Organization: Jinnah Sindh Medical University, Karachi |
| Location: Karachi | Phone:  +92 333 2634 464 |
| Other Investigators :  Eshika  Vandna Devi  JaiRam  Kirshna Chawlla  Sandesh  Vandna Kumari  Jamshaid | Organization: Jinnah Sindh Medical University, Karachi |
| Location; Karachi | Phone:   1. +92 317 3289 854 2. +92 332 7962 289 |

Dear Participant,

I am Dr.Marium Munir Tunio, from Jinnah Sindh Medical University, Karachi. My students and I are currently doing a study on prevalence of Body focused Repetitive University, Karachi. To understand more about this, we would like to invite you to participate in our study. The information below is provided to give you an idea of our research, we request you to please read the information below before you agree to participate:

**1.PURPOSE OF THIS RESEARCH STUDY**

To determine frequency of BFRBs among undergraduate medical students enrolled in MBBS program at Sindh Medical College aged between18-25 years.

**2.PROCEDURES**

As a study participant, you will be asked to fill a questionnaire that will obtain demographic information. This questionnaire will be circulated via online platforms (Email, WhatsApp). This activity is expected to take no more than 5-10 minutes of your time.

3.**POSSIBLE BENEFITS**

As a study participant, you will contribute to important medical research on the prevalence of BFRBs among medical students.

**4.FINANCIAL CONSIDERATIONS**

You are not expected to bear any costs for participation in our research survey, neither are there any monetary benefits you may be entitled to.

**5.AVAILABLE MEDICAL TREATMENT FOR ADVERSE EXPERIENCES**

NA

6. **RIGHT OF REFUSAL TO PARTICIPATE AND WITHDRAWAL**You are free to choose to participate in the study. You may also withdraw any time from the study. You may opt not to answer any question with which you are not comfortable with.
7. **CONFIDENTIALITY**

The information provided by you will remain confidential. Nobody except principal investigator will have access to it. Your identity will also not be disclosed at any time. However, the data may be seen by Ethical review committee and may be published in journal and elsewhere without giving name or disclosing the identity.

**8.PARTICIPATION IN RESEARCH STUDY**

You are free to choose whether or not to participate in this study. There will be no penalty or loss of benefits to which you are otherwise entitled if you choose not to participate. As a participant, you will be informed of any significant new findings that develop from this study.

9.**AVAILABLE SOURCES OF INFORMATION**

If you have any further questions, you may contact Principal Investigator.

**10.AUTHORIZATION**

I have read and understood this assent form. I undertake that the importance and the methods of the research study have been explained to me and I voluntarily participate in it after knowing all the terms and conditions. I understand that my consent does not take away any legal rights in case of negligence or other legal fault of anyone who is involved in this study. I further understand that nothing in this consent form is intended to replace any applicable Federal, state, or local laws.

Name of Participant:

Date:

Signature of Participant: Date:

Signature of Principal Investigator: Dr. Marium Munir Tunio Date:
