## Supplementary material for "Prevalence of Body-focused repetitive behaviours among undergraduate medical students at public sector university karachi": IRB approval

### Jinnah Sindh Medical University (JSMU)
#### Institutional Review Board (IRB)


**Reference No: JSMU/IRB/2023/749** **Date:** 13/11/23

**Dr. Marium Munir Tunio**
Lecturer
Department of Community Medicine
Jinnah Sindh Medical University
Karachi

**Title of Research: PREVALENCE OF BODY FOCUSED REPETITIVE BEHAVIORS AMONG UNDERGRADUATE MEDICAL STUDENTS OF JINNAH SINDH MEDICAL UNIVERSITY, KARACHI, PAKISTAN.**

Respected Madam,

With reference to your email dated 25th October, 2023 regarding response of protocol reference no JSMU/IRB/2023/749 has been reviewed and approved in its 76th meeting dated 28th October, 2023 for a period of one year to conduct this research.

Any change in the protocol or extension in the period of research should be notified to the IRB-JSMU for further process. Interim report on progress of research. should be submitted to IRB from time to time.

Regards,

**Prof. Dr. Huma Shareef**
Co-Chairperson, IRB
Director Research
Department of Research
Jinnah Sindh Medical University

**Prof. Dr. Aasim Ahmad**
Chairman, Institutional Review Board (IRB), JSMU
Co-Chairperson Bioethics Group,
Hon. Senior Lecturer Aga Khan University &
Dean & Chief Nephrologist
The Kidney Centre Post Graduate Training Institute

- The permission or approval from any other government agency is the responsibility of the Principal investigator.


Rafiqui H.J. Shaheed Road, Karachi-75510
; Web: jsmu.edu.pk


Scanned with CamScanner
